# Plasma inflammatory markers and brain white matter microstructure in late middle-aged and older adults

**DOI:** 10.64898/2026.04.20.26351124

**Authors:** Siona Mishra, Corinne Pettigrew, Chidi Ugonna, Nan-kuei Chen, Jennifer B. Frye, Kristian P. Doyle, Lee Ryan, Marilyn Albert, Sara Grace Ho, Abhay Moghekar, Anja Soldan, Elizabeth R. Paitel

## Abstract

Chronic inflammation is a common feature of aging and is observed across various age-related neurodegenerative diseases, including Alzheimer’s disease (AD). It has, however, been challenging to develop measurements of brain structure directly linked to peripheral measures of neuroinflammation. This cross-sectional study examined whether plasma levels of markers related to inflammation are associated with diffusion magnetic resonance imaging (dMRI) measures of white matter microstructure: mean diffusivity (MD) and Neurite Orientation Dispersion and Density Imaging (NODDI) free water fraction (FWF) and orientation dispersion index (ODI). Participants included 457 dementia-free individuals (mean age=63.82, *SD*=7.63). Blood plasma markers related to inflammation included two measures of systemic inflammation, (1) high-sensitivity C-reactive protein (CRP), and (2) a composite of pro-inflammatory cytokines (IL-1α, IL-1β, IL-2, IL-6, IL-8, TNF-α, TNF-β), as well as (3) glial fibrillary acidic protein (GFAP), a measure of astrocytic activation. Higher cytokine composite levels were associated with higher values of all three measures (FWF, ODI, MD) in cerebral white matter, and with higher ODI in the cerebellar peduncles. Higher CRP levels were associated with higher ODI in cerebral and cerebellar white matter. Associations with GFAP were not significant after adjusting for multiple comparisons. Results were consistent after accounting for plasma biomarkers of AD pathology (p-tau_181_/A*β*_42_). Thus, higher levels of peripheral pro-inflammatory markers are associated with white matter microstructure (higher FWF, ODI, and MD), supporting the view that these dMRI-based metrics are sensitive to inflammatory processes. Additionally, the sensitivity of dMRI-based measures to inflammation may differ by inflammatory marker types.

## 1.1. Background

Chronic neuroinflammation is regularly observed in aging (Baylis et al., 2013; Godbout & Johnson, 2009) and is a common feature of various age-related neurodegenerative diseases, including Alzheimer’s disease (AD; Chen et al., 2016; Heneka et al., 2015). Neuroinflammation is an inflammatory response in the brain that is largely mediated by glial cells (i.e., microglia, which are the principal immune cells in the central nervous system, and astrocytes). Activated microglia and astrocytes can release immune mediators, such as cytokines, which may serve protective immune responses in the short-term (for review see Lyman et al., 2014; Shabab et al., 2017). However, chronic neuroinflammation can interfere with neuronal signaling and damage axons, synapses, and myelin, with downstream consequences for white matter integrity (Oestreich & O’Sullivan, 2022; Rodrigue et al., 2019). The availability of measures of brain structure that reflect chronic inflammation would therefore be helpful in understanding brain and cognitive health among older adults.

While levels of systemic inflammation can be easily obtained from blood, measuring inflammation in the brain is difficult due to the highly selective blood-brain barrier (Galea, 2021). Efforts are therefore underway to identify non-contrast magnetic resonance imaging (MRI) measures of neuroinflammation. Although MRI cannot directly image microglia or astrocytes, several MRI metrics have been proposed to reflect brain tissue properties that are closely associated with neuroinflammation (Jelescu & Fieremans, 2023; Oestreich & O’Sullivan, 2022). One such approach is diffusion MRI (dMRI), which measures the diffusion of water in tissue and is used to investigate brain tissue microstructure.

It has been suggested that some dMRI-based metrics are sensitive to neuroinflammation; however, there is relatively little evidence directly linking central or peripheral inflammatory markers to measures of brain tissue microstructure in non-clinical populations (for review see Jelescu & Fieremans, 2023; Oestreich & O’Sullivan, 2022). Two studies among dementia-free older adults using conventional, single-shell diffusion tensor imaging (DTI) did not find significant relationships between levels of pro-inflammatory cytokines in blood (i.e., interleukin 6 [IL-6] and IL-1β) with whole-brain white matter fractional anisotropy (FA; Gu et al., 2017) or mean diffusivity (MD; Boots et al., 2020). In a large sample that was not evaluated for cognitive impairment, higher baseline levels of plasma glial fibrillary acidic factor (GFAP), a marker of astrocytic activation often associated with neuroinflammatory processes, were associated with a longitudinal increase in MD (Huang et al., 2025). A number of studies have examined C-reactive protein (CRP), which is primarily produced by the liver in response to inflammation (Du Clos, 2000; Sproston & Ashworth, 2018). While largely reflective of peripheral inflammation, high levels of CRP may disrupt the blood-brain barrier, thereby contributing to neuroinflammation (Alilou et al., 2024; Hsuchou et al., 2012). Studies among older adults without dementia showed that higher CRP levels or a composite of CRP and TNF-α (Arfanakis et al., 2013), a pro-inflammatory cytokine, were associated with lower white matter FA and/or higher MD (Alilou et al., 2024; Boots et al., 2020; Conole et al., 2021; Miralbell et al., 2012; O’Donovan et al., 2021; Walker et al., 2017; Walker et al., 2018; Wersching et al., 2010). Taken together, these prior studies suggest associations between higher levels of CRP and conventional DTI-based metrics of white matter microstructure (e.g., FA and MD) among dementia-free older adults, though studies examining associations between levels of pro-inflammatory cytokines or GFAP with these metrics are scarce.

Importantly, there is a dearth of research examining associations between markers related to inflammation and more advanced diffusion-based measures from neurite orientation dispersion and density imaging (NODDI). NODDI provides more details about microstructural tissue properties by separating the signal in each voxel into three compartments: within neurites (i.e., axons and dendrites), between neurites, and extracellular, unrestricted (i.e., “free”) water, such as interstitial fluid and cerebrospinal fluid (CSF; Zhang et al., 2012). Animal studies suggest that higher free water fraction (FWF), a NODDI measure reflecting elevated extracellular “free” water, is associated with higher GFAP levels (Garcia-Hernandez et al., 2022; Yi et al., 2019), while higher orientation density index (ODI), a NODDI measure reflecting variability in neurite orientation, is associated with microglial density (Kim et al., 2023; Yi et al., 2019). A recent UK Biobank study found that longitudinal increases in plasma GFAP were associated with decreases in NODDI intracellular volume fraction (ICVF), a measure of neurite density, but not with consistent changes in extracellular volume fraction or orientation dispersion (Huang et al., 2025).

Two previous studies using a bi-tensor model of free water (vs. three-compartment with NODDI) among individuals with AD dementia and other neurodegenerative diseases, reported a positive association between plasma GFAP and FWF (Kim et al., 2025; Sumra et al., 2025). It is unclear if similar associations exist among dementia-free individuals, as the above-mentioned plasma inflammatory markers and white matter NODDI metrics have not been examined within the context of aging.

To address these gaps, the current study examined the cross-sectional associations of multiple plasma measures related to inflammation (a composite of pro-inflammatory cytokines, GFAP, and CRP) with conventional (MD) and NODDI (FWF, ODI) measures of white matter microstructure in a large sample (*N*=457) of middle-aged and older dementia-free adults. Global white matter microstructure was evaluated in both the cerebrum and the cerebellar peduncles, as reports suggest that cerebellar microglia may have specialized immunogenic properties (Dukhinova et al., 2024; Stoessel & Majewska, 2021; Stowell et al., 2018). In exploratory analyses, we also examined whether these associations differed by levels AD pathology (i.e., amyloid and tau protein levels in blood plasma), given that AD pathology accumulation is associated with neuroinflammation (Kamila et al., 2025). We hypothesized that higher levels of inflammatory markers would be associated with higher levels of all of the dMRI measures (MD, FWF, and ODI) in the global cerebral and cerebellar tracts and that these associations may be stronger in individuals with more abnormal AD biomarker levels.

## 2.1. Methods

### 2.1.1. Study Design and Participant Recruitment

The Healthy Minds for Life (HML) study of the Precision Aging® Network (PAN; https://precisionagingnetwork.org/) was designed to investigate factors that predict individual brain health risks with the ultimate goal to optimize brain health across the lifespan and extend the cognitive healthspan (Ryan et al., 2019). This multi-site study includes four sites: the University of Arizona (Tucson, AZ), the University of Miami (Miami, FL), Emory University (Atlanta, GA), and the Johns Hopkins University (Baltimore, MD). Participants met the following inclusion criteria at study entry: 50-79 years of age; self-identify as White, Black, or Hispanic race/ethnicity; English language proficiency; no reported diagnosis of memory loss or dementia; no reported history of psychotic illness including schizophrenia or bipolar disorder; and no contraindications for blood draw or brain MRI. Enrollment for this study began in 2022 and is ongoing. Participants are recruited through multiple approaches, including online methods, in-person recruitment events, and referrals from site-specific research registries. The current analyses included *N* = 457 late middle-aged to older adult participants with diffusion MRI and blood inflammatory marker data. This study was approved by the WIRB-Copernicus Group Institutional Review Board and all participants provided written informed consent.

### 2.1.2. Maghnetic Resonance Imaging Acquisition

MRI scans were acquired on a 3T Phillips Achieva scanner (Eindhoven, The Netherlands) at Johns Hopkins University and on 3T Siemens scanners (Munich, Germany) at the other three sites: Skyra at the University of Arizona, Vida at the University of Miami, and Prisma at Emory University. The multi-modal protocol included a magnetization-prepared rapid gradient echo (MPRAGE) scan used for anatomical reference and image registration (Philips TR = 6500 ms, TE = 2.943ms, FA = 9 degree, resolution 1mm x 1mm x 1mm, size 256 x 256 x 211; Siemens TR = 2300ms, TE =2.98ms, FA = 9 degree, resolution 1mm x 1mm x 1mm, size 240 x 256 x 208). Diffusion-weighted images were acquired using a multi-shell acquisition scheme (resolution 2mm x 2mm x 2mm, TE/TR = 91/3700ms, multiband factor = 3, Philips size 128 x 128 x 69, Siemens size 116 x 116 x 69) with b-values of 500, 1000 and 2000 s/mm^2^. A total of 137 volumes were collected, consisting of 13 non-diffusion-weighted (b0) images and 124 diffusion-weighted images evenly distributed across the three shells. In addition, b0 images with reversed phase-encoding polarity were acquired to estimate and correct susceptibility-induced distortions in the diffusion MRI data.

### 2.1.3. Image Processing

The diffusion-weighted images were pre-processed using QSIPREP version 0.21.4 (Cieslak et al., 2021). Processing included joint eddy current correction, head motion correction and susceptibility distortion correction. White matter bundle segmentation for each participant was performed on the pre-processed QSIPREP outputs using Tractseg (https://github.com/MIC-DKFZ/TractSeg) version 2.8. The qsirecon module of QSIPREP was used to fit a white-matter NODDI model (amico-noddi) using default NODDI parameters. Additional masking was performed using white matter masks from FSL’s FAST algorithm and Freesurfer’s surface pipeline to obtain summaries of FWF, ODI, and MD in global cerebral white matter, which were used to calculate the cerebral white matter measures used in this study. Additionally, tract-level FWF, ODI, and MD values were computed for 38 ROIs, which were used to derive a cerebellar peduncle composite as the average of bilateral inferior, middle, and superior cerebellar peduncle values.

The current study focused on one conventional diffusion measure (i.e., MD) and two NODDI measures (FWF and ODI), as these have been proposed to reflect, at least partially, processes related to neuroinflammation, as described above. MD reflects the average diffusivity of water in all directions (Pierpaoli et al., 1996), providing a measure of overall tissue density and microstructure. Though its biophysical specificity is limited (Jones et al., 2013), lower MD values are generally interpreted as reflecting ‘better’ microstructural integrity, with higher MD related to both greater neurodegeneration (including axonal and synaptic degeneration and loss), as well as edema and inflammatory processes (Jelescu & Fieremans, 2023; Plank et al., 2025; Zhang et al., 2014). From NODDI, higher FWF reflects elevated extracellular “free” water, and is proposed to be a relatively more direct and sensitive metric of neuroinflammation (Kim et al., 2025; Sumra et al., 2025), though it is also related to neurodegeneration (Febo et al., 2020; Kamiya et al., 2020). ODI quantifies variability in neurite orientation, with higher values generally reflecting greater disorganization in the direction of neurites. As ODI and FA metrics tend to be highly anti-correlated and the biological interpretation of ODI is clearer than FA, FA was not analyzed to reduce the number of comparisons. Analyses focused on FWF, ODI, and MD values for the global cerebral white matter as well as a cerebellar peduncle composite. FWF, ODI, and MD values were z-scored within study site prior to averaging.

### 2.1.4. Plasma Inflammatory, GFAP, and AD Biomarker Measures

Plasma was collected via EDTA tubes after an overnight fast; samples were centrifuged at 1500g for 15 minutes at 4°C and stored in −80 in polypropylene vials prior to being shipped on dry ice to the coordinating site (University of Arizona) for processing. Cytokine concentrations were quantified using the MILLIPLEX® MAP Human Cytokine Magnetic Bead Panel (Millipore, Cat# HYCTA60K) according to the manufacturer’s instructions. Plasma samples were thawed on ice and undiluted samples were centrifuged at 10,000 × g for ten minutes to remove debris. Standards and quality controls were reconstituted and serially diluted as specified by the manufacturer. The assay was performed on a Luminex® LX200 instrument, equipped with xPONENT® software. A minimum of 100 beads per analyte were collected per well. Standard curves were generated using a 5-parameter logistic (5-PL) curve-fitting algorithm using Millipore’s Belysa Analysis Software. Cytokine concentrations were calculated by interpolation from the standard curves and expressed in pg/mL. Samples with values below the lower limit of detection (LLOD) were reported as the minimum detectable concentration reported by Millipore divided by two.

Inflammatory markers of interest included: IL-1α, IL-1 β, IL-2, IL-6, IL-8, TNF-α, and TNF-β. Given the lack of *a priori* hypotheses regarding specific cytokines, markers were z-score transformed and then averaged to create a pro-inflammatory composite score. Preliminary analyses showed that these measures tended to show weak positive correlations with one another, supporting the approach of combining them into a single measure. While less specific than the individual markers, this pro-inflammatory cytokine composite may be a more sensitive measure of overall inflammation. High-sensitivity CRP was measured by Sonora Quest Laboratories using a chemiluminescent immunoassay. CRP was analyzed separate from the pro-inflammatory cytokine composite given biological differences in the production, time course, and function. CRP values were log-transformed to reduce skewness and then z-score normalized.

Plasma samples were analyzed for GFAP, p-tau_181_, and Aβ_42_ using the Single Molecule Array (Simoa) Neurology 4-Plex E, with p-tau_181_ assays on the HD-X instrument (Quanterix Corporation). Quanterix Simoa is an ultrasensitive assay that uses monoclonal antibodies and calibrators to allow for detection at very low concentrations. All assays were run in duplicate, and the mean value of the duplicate measurements was used for all analyses. Intra-assay CVs for all analytes were less than 10% and inter-assays CVs for all analytes were less than 20% for a native plasma sample run across all plates in addition to high and low kit controls supplied by the manufacturer. GFAP scores were log-transformed to reduce skewness and then z-scored. AD biomarker levels were quantified using the ratio of p-tau_181_/Aβ_42_ (Fowler et al., 2022), which was z-scored prior to analysis. The percentage of participants with abnormal levels of AD biomarkers (11% for A*β*_42_/A*β*_40_, 4.49% for p-tau_181_) were determined based on A*β* clinical cut-points (Quaresima et al., 2024) and company reported p-tau norms.

### 2.1.5. Cognitive Assessment

Participants completed a short cognitive battery during their visit that included the Montreal Cognitive Assessment (MoCA), a screener providing a measure of global cognitive performance (Nasreddine et al., 2005). A previously validated MoCA cut-point of 24 was applied for sensitivity analyses to examine relationships among participants without cognitive impairment (Malek-Ahmadi & Nikkhahmanesh, 2024). A meta-analysis that evaluated the MoCA for detection of mild cognitive impairment demonstrated that a cut-point of 24 has good sensitivity (80%, range = 67–88%) and specificity 84% (range 75–90%; Islam et al., 2023). Values for dementia are reported to be even higher, with a very large, nation-wide study showing sensitivity 95.3% and specificity 92.67% (Ismail et al., 2025). Additional cognitive testing and surveys (not included in these analyses) were completed in-person as well as online via the MindCrowd.org website.

### 2.1.6. Statistical Analysis

Multiple linear regression models were used to examine the cross-sectional associations of markers related to inflammation (i.e., CRP, pro-inflammatory cytokine composite, and GFAP, the primary predictors of interest, evaluated in separate models) with the global cerebral and cerebellar peduncle white matter microstructure measures as outcomes (using separate models for FWF, ODI, and MD). All models covaried age, sex, and study site. A set of exploratory follow-up models examined whether the observed associations between inflammatory levels and diffusion measures differed by AD biomarker levels by adding terms for p-tau_181_/Aβ_42_ and the inflammation*p-tau_181_/Aβ_42_ interaction to the primary models, as well as body mass index (BMI). To address concerns regarding multiple comparisons, a False Discovery Rate (FDR) correction was applied for all analyses, adjusting for six comparisons (i.e., cerebral and cerebellar MD, FWF, and ODI metrics) separately for the three plasma inflammatory measures (Benjamini & Hochberg, 1995).

A sensitivity analysis examined whether the results from primary models changed when excluding participants with a MoCA score below 24, which may be indicative of cognitive impairment (Malek-Ahmadi & Nikkhahmanesh, 2024). A second sensitivity analysis examined whether results changed when covarying BMI and a summary vascular risk score (see Supplementary Table 2 for details), given vascular health may influence both inflammatory markers and white matter microstructure (e.g., Jacobs et al., 2013; Kennedy & Raz, 2009; Moyse et al., 2022; Williams et al., 2019).

## 3.1. Results

### 3.1.1. Descriptive statistics

Descriptive statistics for the 457 participants included in the analyses are shown in Table 1. On average, participants were 63.8 years old, predominantly female (69%), and 37% from racial or ethnic groups traditionally underrepresented in research. The two systemic inflammatory measures were significantly correlated with one other (CRP-cytokines: *r* = 0.15, *p* < 0.01) but not with GFAP (CRP-GFAP: *r* = 0.04, *p* = 0.44, cytokines-GFAP: *r* = 0.08, *p* = 0.07). The correlations among dMRI metrics ranged from non-significant to moderate (*r* = 0.02 to 0.68, *p* = 0.66 to < 0.001, see Supplemental Table 4).

**Table 1.** Descriptive statistics for total sample.

|  | N=457 |
| --- | --- |
| Age | 63.82 (7.63) |
| Sex, *n* (%) female | 314 (68.7%) |
| Ethnicity (% Hispanic/Latino) | 76 (17%) |
| Race |  |
| Black or African American, *n* (%) | 91, 19.91% |
| White, *n* (%) | 356, 77.90% |
| Other/multi-racial, *n* (%) | 10, 2.19% |
| Education level |  |
| High School diploma, *n* (%) | 31 (6.8%) |
| Some college or less, *n* (%) | 89 (19.5%) |
| Undergraduate degree, *n* (%) | 173 (37.9%) |
| Post-graduate degree, *n* (%) | 164 (35.9%) |
| MoCA | 26.31 (2.64) |
| MoCA < 24, *n* (%) | 71 (16%) |
| CRP | 2.54 (4.37) |
| Cytokine composite | 0.00 (0.71) |
| GFAP | 135.00 (76.70) |
| p-tau<sub>181</sub>/Aβ<sub>42</sub> | 3.81 (2.10) |
| BMI | 28.00 (6.00) |

Note: MoCA = Montreal Cognitive Assessment total score. Numbers indicate mean (SD), unless otherwise noted.

### 3.1.2. Associations of inflammatory markers with white matter microstructure measures

In the primary models, higher cytokine composite levels were associated with higher FWF, ODI, and MD in cerebral white matter (*p*s < 0.05), as well as higher ODI in the cerebellar peduncle composite (*p* = 0.001; see Table 2, Figure 1). Higher CRP levels were associated with higher cerebral (*p* < 0.05) and cerebellar (*p* < 0.01) ODI. Higher GFAP levels were associated with lower cerebral FWF (*p* < 0.05), which did not remain significant following correction for multiple comparisons.

**Figure 1.**
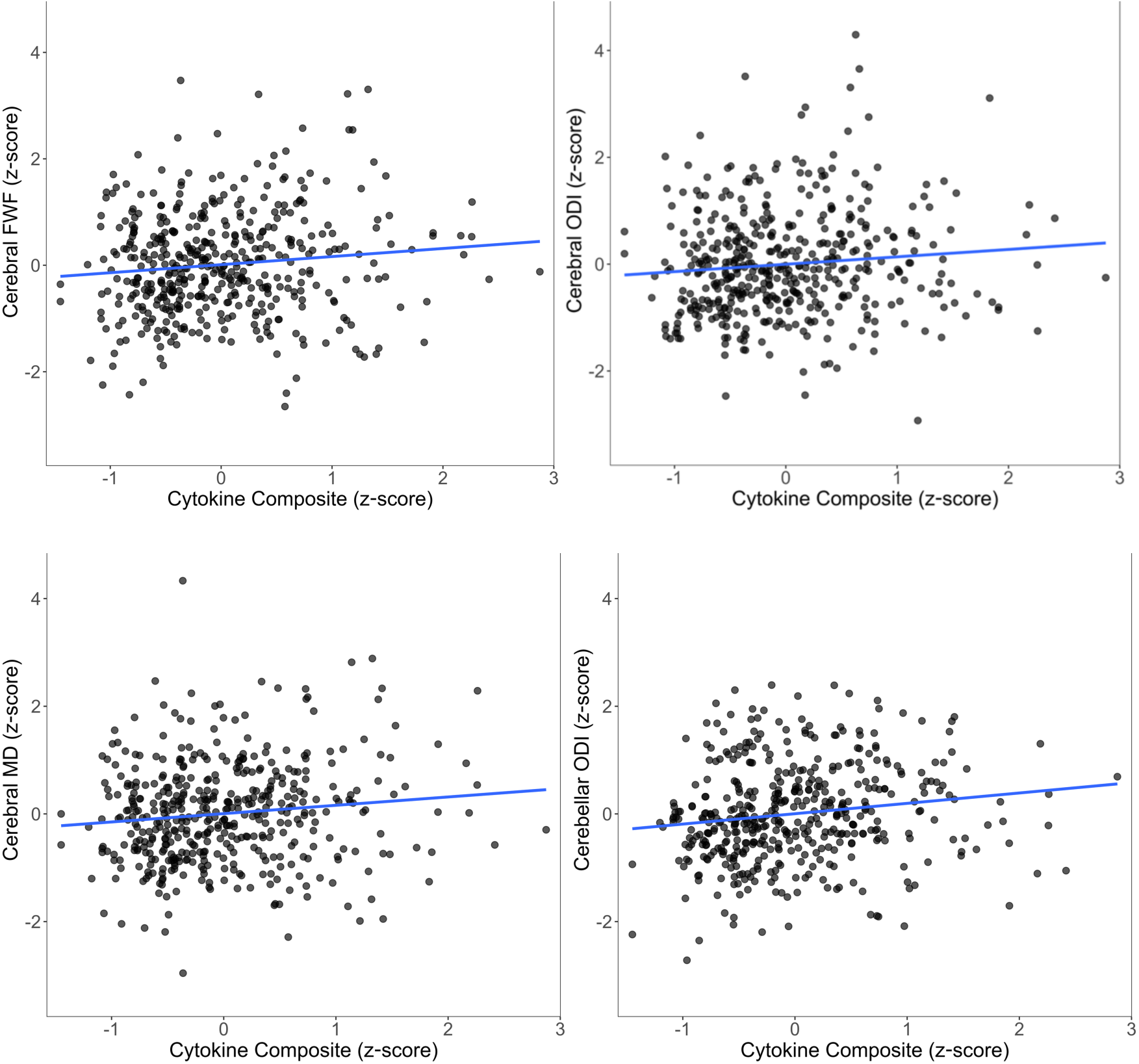
Higher levels of a pro-inflammatory cytokine composite (including IL-1α, IL-1 β, IL-2, IL-6, IL-8, TNF-α, and TNF-β) are associated with higher cerebral white matter FWF (top left), ODI (top right), MD (bottom left), and cerebellar peduncle ODI (bottom right). DTI metrics are shown residualizing age, sex, and study site. Similar patterns were evident for CRP with cerebral and cerebellar ODI (figures not shown).

**Table 2.** Results from multiple linear regression models assessing the associations of systemic inflammatory and GFAP levels with white matter microstructure, covarying age, sex, and study site.

| **Cerebral** |  | **Coeff** | **SE** | **t** | **p** | **Cerebellar** |  | **Coeff** | **SE** | **t** | **p** |
| --- | --- | --- | --- | --- | --- | --- | --- | --- | --- | --- | --- |
| <u>Cytokine Composite</u> |  |  |  |  |  | <u>Cytokine Composite</u> |  |  |  |  |  |
|  | FWF | **0.158** | **0.065** | **2.418** | **0.016\*** |  | FWF | 0.014 | 0.058 | 0.245 | 0.807 |
|  | ODI | **0.140** | **0.066** | **2.131** | **0.034** |  | ODI | **0.193** | **0.060** | **3.241** | **0.001\*** |
|  | MD | **0.155** | **0.062** | **2.508** | **0.013\*** |  | MD | 0.004 | 0.057 | 0.07 | 0.944 |
| <u>CRP</u> |  |  |  |  |  | <u>CRP</u> |  |  |  |  |  |
|  | FWF | -0.007 | 0.048 | -0.146 | 0.884 |  | FWF | -0.030 | 0.031 | -0.948 | 0.344 |
|  | ODI | **0.120** | **0.048** | **2.496** | **0.013\*** |  | ODI | **0.126** | **0.043** | **2.890** | **0.004\*** |
|  | MD | 0.014 | 0.046 | 0.308 | 0.758 |  | MD | 0.004 | 0.041 | 0.105 | 0.917 |
| <u>GFAP</u> |  |  |  |  |  | <u>GFAP</u> |  |  |  |  |  |
|  | FWF | **-0.127** | **0.055** | **-2.287** | **0.023** |  | FWF | -0.096 | 0.049 | -1.957 | 0.051 |
|  | ODI | -0.089 | 0.055 | -1.602 | 0.110 |  | ODI | -0.098 | 0.051 | -1.924 | 0.055 |
|  | MD | 0.090 | 0.052 | 1.708 | 0.088 |  | MD | 0.065 | 0.048 | 1.365 | 0.173 |

Note. **Bold**, un-corrected $p < 0.05$ . \*Effect remains significant after applying FDR correction for multiple comparisons (six comparisons, FDR-adjusted $p$ value $< 0.05$ ). coeff = coefficient; FWF = free water fraction, MD = mean diffusivity, ODI = orientation dispersion index, SE = standard error.

For model covariates, older age was associated with higher cerebral FWF, higher cerebral ODI (in CRP model only), and higher cerebral and cerebellar MD (*p*s < 0.05). For sex, cerebral ODI values were higher in men (*p*s < 0.01); men also had higher cerebellar ODI in the CRP model (*p*s < 0.05). Study site was not a significant covariate in any model (*p*s > 0.623).

In the exploratory follow-up analyses including interactions with AD biomarker levels, there was one significant pro-inflammatory cytokine composite*p-tau_181_/A*β*_42_ interaction for cerebellar MD (*p* = 0.024), although this did not remain significant following FDR adjustment for multiple comparisons. All other interaction terms were non-significant (*p*s > .073, data not shown). When the non-significant interaction terms were removed, there were no significant associations between p-tau_181_/A*β*_42_ and white matter microstructure (*p*s > .064, data not shown).

In the sensitivity analyses, the results of the primary models were consistent when excluding participants with a MoCA score below 24 (Supplementary Table 1). When BMI and a vascular risk summary score were included as additional covariates, the cytokine composite and GFAP results were comparable, although associations for CRP were no longer significant (Supplementary Table 2). Supplementary Table 3 shows results using the individual cytokines, rather than the cytokine composite score. Overall, IL1α, IL6, and IL8 had the most robust and consistent associations with the white matter microstructure metrics.

## 4.1. Discussion

The goal of this study was to examine cross-sectional associations between peripheral measures of inflammation and measures of white matter microstructure based on dMRI. In late middle-aged and older adults, higher levels of a pro-inflammatory cytokine composite were associated with higher levels of all three dMRI measures examined (MD, FWF, and ODI) in cerebral white matter, whereas the cerebellar peduncle composite was only associated with higher levels of ODI. Higher CRP levels were associated with higher cerebral and cerebellar ODI. Higher GFAP was associated with lower cerebral FWF, which did not remain significant after adjusting for multiple comparisons. These patterns were evident even after accounting for biomarkers of AD pathology. Taken together, these findings suggest that systemic inflammation is linked to alterations in both cerebral and cerebellar white matter microstructure integrity, and that these diffusion measures partially reflect inflammatory processes. By comparison, global cerebral and cerebellar white matter microstructure may be less sensitive to alterations in plasma GFAP, a measure of astrocytic activation.

The most robust relationships with white matter microstructure were observed for the composite of pro-inflammatory cytokines. The few prior studies that examined plasma cytokine levels in relationship to dMRI measures of brain microstructure among dementia-free older adults did not find significant associations for the individual cytokines IL-6 or IL-1β with FA (Gu et al., 2017) or MD (Boots et al., 2020). In contrast, we found that higher cytokine composite levels were associated with higher cerebral MD. Our positive findings may be due to the use of a composite of various pro-inflammatory cytokines, which may provide greater sensitivity in assessing inflammation, as it captures a greater array of inflammatory responses.

The current study further expands on existing research by showing higher levels of the pro-inflammatory composite were associated with higher dMRI values based on NODDI, with higher FWF in cerebral white matter and higher ODI in both cerebral and cerebellar peduncle tracts. Thus, this newer multi-shell acquisition approach to the assessment of white matter microstructure may be particularly valuable for assessing relationships with inflammation.

The study also found that higher levels of CRP were associated with higher ODI in both cerebral white matter and the cerebellar peduncle composite. These findings expand upon previous studies that have primarily analyzed CRP in relationship to conventional, single-shell diffusion metrics (Boots et al., 2020; Conole et al., 2021; Miralbell et al., 2012; O’Donovan et al., 2021; Walker et al., 2017; Walker et al., 2018; Wersching et al., 2010), and suggest that higher CRP may be related to more dispersed or disorganized neurites among late middle-aged and older adults. However, these results did not remain significant when covarying vascular risk factors, suggesting that relationships between CRP and dMRI-based measures of brain microstructure may be confounded by levels of vascular risk. Additionally, unlike some previous longitudinal studies (Conole et al., 2021; O’Donovan et al., 2021; Walker et al., 2017; Walker et al., 2018), we did not find significant associations between CRP and MD. These differences may be attributable to the cross-sectional nature of the current study and sample characteristics, such as vascular risk, with most of the previous studies being comprised of participants with greater vascular and medical risk factors. Our results therefore highlight the importance of controlling for BMI and other vascular risk factors when investigating plasma markers related to inflammation and measures of brain microstructure. Future studies directly evaluating whether associations between DTI metrics and CRP differ by level of vascular risk may provide valuable insight.

To our knowledge, only one previous study has examined relationships between plasma GFAP and white matter dMRI metrics in dementia-free participants, finding that higher baseline GFAP was associated with a longitudinal increase in MD, but reporting no consistent associations with NODDI orientation dispersion or extracellular water diffusion (Huang et al., 2025). The present study found that higher GFAP was associated with lower cerebral FWF, but this effect did not remain significant after adjusting for multiple comparisons, which is consistent with the lack of robust relevant NODDI effects reported by Huang and colleagues. The lack of significant associations with MD in the current analyses may be attributable to sample characteristics or the cross-sectional nature of the present study. In two studies of older participants across the AD spectrum, higher GFAP levels were associated with lower FA and higher MD (Bettcher et al., 2021; Kim et al., 2025), as well as higher free-water fractional volume using bi-tensor free water imaging (Kim et al., 2025). Thus, future longitudinal research is needed to determine whether associations between GFAP and dMRI are more robust in later disease stages, which are characterized by higher levels of neurodegeneration and pathology.

Taken together, these patterns demonstrate the importance of examining multiple markers of systemic inflammation and astrocytic activation, which may capture different aspects of the inflammatory response. CRP is produced in response to inflammation, including the presence and increase of pro-inflammatory cytokines (Du Clos, 2000; Sproston & Ashworth, 2018). While CRP and cytokine levels are general, non-specific markers of inflammation, GFAP is thought to primarily reflect astrocytic activation, lending it more specificity to brain and central nervous system inflammation (Eng & Ghirnikar, 1994). However, GFAP levels measured in plasma (vs. cerebrospinal fluid) may reflect a mix of other neurodegenerative or inflammatory processes, particularly in the absence of blood-brain barrier damage (Youn et al., 2025). Future studies examining GFAP and cytokine levels in CSF are therefore needed to determine whether they are similarly related to microstructural properties.

Regarding the diffusion metrics, the current findings demonstrated that higher levels of all of the measures examined (MD, FWF, and ODI) were associated with higher levels of a pro-inflammatory cytokine composite. In contrast, CRP and GFAP were each associated with one metric derived from the multi-shell acquisition, but not the standard metric of MD. Thus, while NODDI metrics (e.g., FWF and ODI) may provide more sensitivity for some markers related to inflammation, MD, which can be obtained using a conventional, single-shell acquisition protocol is still informative. This is consistent with suggestions that all three metrics may reflect neuroinflammation to some degree (Jelescu & Fieremans, 2023; Kim et al., 2025; Kim et al., 2023; Plank et al., 2025; Sumra et al., 2025; Yi et al., 2019). Future studies are needed to replicate these findings, and to further explore the mechanisms underlying the reported associations.

In contrast to the cerebral white matter, relationships between microstructure of the cerebellar peduncles and inflammatory markers were specific to ODI metrics, and not significant for FWF or MD. Specifically, higher CRP and pro-inflammatory cytokines were both associated with higher ODI in a composite of the cerebellar peduncles, which are the white matter tracts that connect the cerebellum with the rest of the brain and spinal cord. Because the cerebellum is often omitted from neuroimaging research, this expands prior work. Although the cerebellum is important for cognitive functioning (Buckner, 2013; Schmahmann, 2019; Schmahmann et al., 2019; Stoodley, 2012) and changes with aging (Arleo et al., 2024; Bernard, 2022) and in various neurodegenerative diseases (Gellersen et al., 2021; Gellersen et al., 2017; Hoxha et al., 2018; Liu et al., 2024; Schmahmann, 2016), relatively little is known about markers of inflammation in the human cerebellum. Of possible relevance, higher ODI has also been suggested to correspond with microglial density in animal models (Kim et al., 2023; Yi et al., 2019). Animal models further suggest that activated cerebellar microglia release pro-inflammatory cytokines, including TNFα, IL-1β, and IL-6, and that the latter two cytokines mediate cerebellar neuronal activity (Dukhinova et al., 2024; Gyengesi et al., 2019; Yamamoto et al., 2019). Additionally, cerebellar microglia have unique features that have led some to consider them a separate subset compared to microglia in the cerebral cortex, including distinct morphology, distribution, functional roles, and molecular structure (Dukhinova et al., 2024; Stoessel & Majewska, 2021; Stowell et al., 2018). These differences may contribute to the unique associations of cytokine and CRP levels with cerebellar ODI (but not FWF or MD) in the present study.

In this study, the observed relationships between inflammatory markers with white matter microstructure were largely independent of biomarkers of AD pathology. While some previous studies have reported relationships between diffusion metrics and AD biomarkers in cognitively unimpaired samples and theorized that the patterns may be attributable to inflammation, these previous studies did not include measures of inflammation (Benitez et al., 2022; Collij et al., 2021; Dong et al., 2020; Hoy et al., 2017; Sun et al., 2024). A number of factors may have contributed to the lack of associations of AD biomarkers with GFAP and the inflammatory measures in the current study. Notably, our sample included many middle-aged participants and was largely cognitively unimpaired. This resulted in a small percentage of participants with abnormal levels of AD biomarkers (11% for A*β*_42_/A*β*_40_, 4.49% for p-tau_181_). Interactions between AD biomarkers and measures of inflammation are likely to be more evident in samples that have greater AD pathology accumulation, such as those in the preclinical or early symptomatic stages of AD.

This study has several strengths, including a large, multi-site cohort of participants with multiple inflammation-related plasma measures, multi-compartment diffusion metrics, and AD biomarker measures. Limitations should also be noted. First, this was a cross-sectional, observational study. We therefore cannot infer directionality of the relationships between these plasma and diffusion measures, or causal relationships. Second, future studies with participants who have higher levels of AD pathology are necessary to evaluate potential interactions between inflammation and AD pathology. Third, the present study used plasma measures of inflammation. It is uncertain to what degree these peripheral measures reflect neuroinflammation (e.g., Youn et al., 2025). Thus, future studies are needed examining measures of inflammation in CSF and their associations with dMRI metrics. Finally, the current analyses investigated global cerebral white matter tracts and the cerebellar peduncles. Future analyses interrogating regionally specific white matter microstructure may reveal patterns that are unique to different regions and tracts.

## Data Availability

Data used in these analyses are available through standard application procedures described on the PAN website (precisionagingnetwork.org).

## Acknowledgements

We thank the Precision Aging® Network for their contribution to the research. The members of the Precision Aging® Network are A. Aldabergenova, A. Bilgin, A. Bonfitto, A. Box, A. Dolby, A. Feal Rodriguez, A. Glinka, A. Moghekar, A. Sidhu, A. Sokan, A. Soldan, A.I. Levey, A.J.B. Lee, B. Aimagamabetova, B. Levin, B. Najafi, B. Nursal, B. Stark, B.J. LaFleur, C.A. Barnes, C.A. Pettigrew, C. Anderson, C. Babbitt, C. Camargo, C. Carrasco, C.H. Na, C. Ugonna, C. Ye, C.S. Mitchell, D. Cabral, D. Chambers, D. Coon, D. Kartchner, D. Metz, D. Sama-Borbon, E. Burrows, E.M. Sternberg, E. Paitel, F. Taguinod, G. Leito, G. Pfaff, H. Venkatachalam, J. Don, J. Pekar, J.R. Runyon, J. Simon, J. Sloan, J. Xie, J. Zhou, J.B. Frye, J.G. Varelo Saboria, J.J Lah, K.A. Delgado, K. Ellingson, K. Johnson, K. Norton, K.P. Doyle, K. Sanders, L.F. Gladulich, L. Gossa, L. Ryan, L. White, M. Albert, M. Dehghan-Rouzi, M. Fan, M. Hay, M. Johnson, M. Lee, M. M. Crespo, M.R. Mehl, M. Modjeski, M. Naymik, M. Swartzlander, M. Taylor John, M.D. De Both, M.J. Huentelman, N. Bhadra, N. Merchant, N. Schork, NK. Chen, P. F. Worley, P. Pattany, R.D. Brinton, R. Tandon, S. Beres, S. Callahan, S. Fox-Rosellini, S. Han, S. Hoscheidt, S. Matijevic, S. Merritt, S. Scott, S. Sharma, S. Sockanathan, S. Soto, S.-E. Roh, S.-Y. Kim, T. James, T. Nuno, T. Rundek, T. Smith, T. Trouard, T. Yuhas, T.K. Zepeda, V.D. Calhoun, V.M. Dotson, V. Pfeifer, W. Degnan III, X. Sun, Y. Yang, Y.F. Bolla, and Z. Chen.

