## Supplemental Materials for "Plasma inflammatory markers and brain white matter microstructure in late middle-aged and older adults"

**Sensitivity analyses excluding participants with a MoCA total score <24.**

Sensitivity analyses were conducted excluding *n* = 71 participants with a MoCA total score below 24 (Malek-Ahmadi & Nikkhahmanesh, 2024), resulting in a sample of *n* = 386 participants (mean age = 63.4 (*SD* = 7.4), 68.7% female). Supplemental Table 1 shows results from analyses evaluating the association between markers related to inflammation with white matter microstructure metrics. The pattern of results was consistent with those reported in the primary models for the full sample.

*Supplemental Table 1.* Results from multiple linear regression models assessing the associations of white matter microstructure with age, sex, study site, and systemic inflammatory and GFAP levels, excluding participants with a MoCA score below 24.

| **Cerebral** | Coeff | *SE* | *t* | *p* | **Cerebellar** | Coeff | *SE* | *t* | *p* |
|---|---|---|---|---|---|---|---|---|---|
| Cytokines | | | | | Cytokines | | | | |
| FWF | **0.168** | **0.070** | **2.386** | **0.018** | FWF | -0.011 | 0.061 | -0.182 | 0.855 |
| ODI | **0.139** | **0.069** | **2.013** | **0.045** | ODI | **0.180** | **0.063** | **2.851** | **0.005** |
| MD | **0.135** | **0.066** | **2.037** | **0.042** | MD | -0.026 | 0.057 | -0.467 | 0.641 |
| CRP | | | | | CRP | | | | |
| FWF | -0.016 | 0.055 | -0.292 | 0.771 | FWF | -0.012 | 0.048 | -0.245 | 0.806 |
| ODI | **0.120** | **0.054** | **2.218** | **0.027** | ODI | **0.125** | **0.049** | **2.543** | **0.011** |
| MD | -0.023 | 0.052 | -0.449 | 0.654 | MD | -0.047 | 0.044 | -1.066 | 0.287 |
| GFAP | | | | | GFAP | | | | |
| FWF | -0.111 | 0.063 | -1.765 | 0.078 | FWF | -0.103 | 0.054 | -1.906 | 0.057 |
| ODI | -0.080 | 0.062 | -1.295 | 0.196 | ODI | -0.058 | 0.057 | -1.012 | 0.312 |
| MD | 0.110 | 0.059 | 1.851 | 0.065 | MD | 0.068 | 0.051 | 1.350 | 0.178 |

**Sensitivity analyses covarying vascular risk and BMI.**

Sensitivity analyses examined whether covarying BMI and other vascular risk factors impacted the relationships between white matter microstructure and markers related to inflammation. These models included a composite vascular risk score and BMI as additional model covariates. The composite vascular risk score reflected the sum of four dichotomous vascular risk factors, based on prior publications (Gottesman et al., 2017; Pettigrew et al., 2020; Soldan et al., 2020): hypertension, high cholesterol, diabetes, and smoking. BMI was included as a separate continuous covariate because previous studies have suggested that BMI in particular my influence plasma levels of inflammatory markers (Blüher et al., 2005; Guldiken et al., 2007). Supplemental Table 2 shows that the CRP associations were no longer significant with these vascular risk variables included as additional model covariates, but across the remaining models, the pattern of results was consistent with the primary models.

*Supplemental Table 2.* Results from multiple linear regression models assessing the associations with white matter microstructure with age, sex, study site, systemic inflammatory and GFAP levels, BMI and vascular risk

| **Cerebral** | Coeff | *SE* | *t* | *p* | **Cerebellar** | Coeff | *SE* | *t* | *p* |
|---|---|---|---|---|---|---|---|---|---|
| Cytokines | | | | | Cytokines | | | | |
| FWF | **0.170** | **0.066** | **2.576** | **0.010** | FWF | 0.022 | 0.059 | 0.380 | 0.704 |
| ODI | 0.101 | 0.065 | 1.551 | 0.122 | ODI | **0.147** | **0.059** | **2.520** | **0.012** |
| MD | **0.163** | **0.062** | **2.635** | **0.009** | MD | 0.020 | 0.057 | 0.342 | 0.732 |
| CRP | | | | | CRP | | | | |
| FWF | 0.003 | 0.054 | 0.056 | 0.955 | FWF | 0.049 | 0.048 | 1.023 | 0.307 |
| ODI | 0.020 | 0.053 | 0.383 | 0.702 | ODI | 0.000 | 0.000 | 0.662 | 0.508 |
| MD | 0.015 | 0.051 | 0.291 | 0.771 | MD | 0.046 | 0.047 | 0.992 | 0.322 |
| GFAP | | | | | GFAP | | | | |
| FWF | **-0.132** | **0.056** | **-2.375** | **0.018** | FWF | **-0.104** | **0.049** | **-2.107** | **0.036** |
| ODI | -0.060 | 0.055 | -1.086 | 0.278 | ODI | -0.062 | 0.050 | -1.261 | 0.208 |
| MD | 0.091 | 0.052 | 1.743 | 0.082 | MD | 0.054 | 0.048 | 1.125 | 0.261 |

**Models with individual cytokines**

Supplemental Table 3 shows results from analyses evaluating the association between individual pro-inflammatory cytokines, rather than the inflammatory composite used in the primary analyses, with white matter microstructure metrics for global cerebral and cerebellar peduncle white matter tracts. There was one missing value for IL-1β, IL-2, and IL-6, resulting in sample sizes of $n$=456 for models with those markers. Overall, IL1a, IL6, and IL8 had the most robust and consistent associations with DTI metrics.

*Supplemental Table 3.* Results from multiple linear regression models assessing the associations of white matter microstructure with age, sex, study site, and individual pro-inflammatory cytokine levels

| **Cerebral** | Coeff | *SE* | *t* | *p* | **Cerebellar** | Coeff | *SE* | *t* | *p* |
|---|---|---|---|---|---|---|---|---|---|
| IL1a | | | | | IL1a | | | | |
| FWF | **0.019** | **0.046** | **2.343** | **0.020** | FWF | 0.029 | 0.041 | 0.693 | 0.489 |
| ODI | 0.024 | 0.047 | 0.520 | 0.604 | ODI | **0.096** | **0.042** | **2.259** | **0.024** |
| MD | **0.115** | **0.044** | **2.630** | **0.009** | MD | 0.027 | 0.040 | 0.677 | 0.499 |
| IL1b | | | | | IL1b | | | | |
| FWF | 0.089 | 0.047 | 1.910 | 0.057 | FWF | 0.023 | 0.042 | 0.552 | 0.581 |
| ODI | 0.012 | 0.047 | 0.247 | 0.805 | ODI | 0.062 | 0.043 | 1.442 | 0.150 |
| MD | 0.076 | 0.044 | 1.710 | 0.088 | MD | 0.036 | 0.040 | 0.901 | 0.368 |
| IL2 | | | | | IL2 | | | | |
| FWF | 0.081 | 0.047 | 1.725 | 0.085 | FWF | -0.008 | 0.042 | -0.205 | 0.838 |
| ODI | 0.045 | 0.047 | 0.971 | 0.332 | ODI | 0.048 | 0.043 | 1.114 | 0.266 |
| MD | **0.087** | **0.044** | **1.964** | **0.050** | MD | 0.001 | 0.040 | 0.019 | 0.985 |
| IL6 | | | | | IL6 | | | | |
| FWF | 0.036 | 0.047 | 0.768 | 0.443 | FWF | 0.011 | 0.041 | 0.267 | 0.790 |
| ODI | **0.139** | **0.046** | **3.013** | **0.003** | ODI | **0.179** | **0.042** | **4.280** | **<0.001** |
| MD | **0.106** | **0.044** | **2.423** | **0.016** | MD | 0.000 | 0.000 | -0.012 | 0.990 |
| IL8 | | | | | IL8 | | | | |
| FWF | **0.108** | **0.047** | **2.311** | **0.021** | FWF | -0.015 | 0.042 | -0.365 | 0.715 |
| ODI | **0.118** | **0.047** | **2.529** | **0.012** | ODI | **0.126** | **0.043** | **2.943** | **0.003** |
| MD | 0.084 | 0.044 | 1.893 | 0.059 | MD | -0.066 | 0.041 | -1.627 | 0.104 |
| TNFa | | | | | TNFa | | | | |
| FWF | 0.062 | 0.047 | 1.322 | 0.187 | FWF | 0.007 | 0.041 | 0.162 | 0.872 |
| ODI | 0.101 | 0.046 | 2.177 | 0.300 | ODI | **0.107** | **0.042** | **2.519** | **0.012** |
| MD | 0.061 | 0.044 | 1.390 | 0.165 | MD | 0.043 | 0.040 | 1.071 | 0.285 |
| TNFb | | | | | TNFb | | | | |
| FWF | 0.074 | 0.047 | 1.580 | 0.115 | FWF | 0.005 | 0.042 | 0.115 | 0.909 |
| ODI | 0.053 | 0.047 | 1.120 | 0.263 | ODI | 0.064 | 0.043 | 1.477 | 0.140 |
| MD | 0.019 | 0.045 | 0.426 | 0.670 | MD | -0.026 | 0.041 | -0.643 | 0.521 |

### Correlations among DWI metrics

Supplemental Table 4 shows the correlations among DWI metrics. The strongest correlations were between FWF and MD, both in cerebral white matter ($r = 0.614$) and in the cerebellar peduncle composite ($r = 0.681$). The weakest relationships were observed between cerebral ODI and both cerebral FWF ($r = 0.022$) and cerebral MD ($r = 0.070$).

*Supplementary Table 4*. Pearson correlation matrix showing relationships between DWI metrics.

| | 1. | 2. | 3. | 4. | 5. | 6. |
|---|---|---|---|---|---|---|
| 1. Cerebral FWF | - | | | | | |
| 2. Cerebellar FWF | 0.401*** | - | | | | |
| 3. Cerebral ODI | 0.022 | -0.104* | - | | | |
| 4. Cerebellar ODI | 0.130** | -0.109* | 0.329*** | - | | |
| 5. Cerebral MD | 0.614*** | 0.216*** | 0.070 | 0.130** | - | |
| 6. Cerebellar MD | 0.158*** | 0.681*** | -0.111* | -0.294*** | 0.363*** | - |

Note: ***p<.001, **p<.01, *p<.05.
